# Physical Activity Initiation and Incident Mobility Difficulty Among Older Adults: A Sequential Target Trial Emulation

**DOI:** 10.64898/2026.10.14.26365469

**Authors:** Jingchen Liang

## Abstract

Although physical activity is associated with better physical functioning, the effects of initiating regular physical activity among older adults with persistent inactivity remain unclear. We conducted a sequential target trial emulation using Health and Retirement Study (HRS) data to estimate the effect of physical activity initiation versus noninitiation on incident mobility difficulty. Participants were included in the analysis if they were aged ≥65 years at the previous wave, reported no regular physical activity at the previous two waves, and had no mobility, activities of daily living (ADL), or instrumental activities of daily living (IADL) difficulties at baseline. Stabilized inverse probability of treatment weighting was applied to adjust for measured confounding. Weighted cumulative incidence was then estimated with death treated as a competing event, and risk differences were calculated at approximately 2, 4, and 6 years. At 2 years, the cumulative incidence of mobility difficulty was 32.75% among initiators and 39.73% among noninitiators, with a risk difference of −6.98 percentage points (95% CI, −13.11 to −0.88). At 4 and 6 years, risk differences were −2.90 percentage points (95% CI, −9.77 to 4.29) and −5.44 percentage points (95% CI, −12.27 to 1.99), respectively. Initiating regular moderate or vigorous physical activity was associated with a lower risk of incident mobility difficulty at approximately 2 years among older adults with persistent inactivity. This difference was less evident at later follow-up, and no clear differences were observed for incident ADL or IADL difficulty.

## INTRODUCTION

Physical inactivity is an important public health concern among older adults. Activity levels tend to decline with age, and a substantial proportion of older adults do not meet recommended physical activity levels (1,2). Physical inactivity has been associated with a higher risk of functional limitation, disability, and loss of independence in later life (3,4). Mobility is particularly important for maintaining independence and is closely related to subsequent disability, morbidity, and mortality among older adults (5).

Regular physical activity may help preserve physical function and prevent mobility disability. In the Lifestyle Interventions and Independence for Elders (LIFE) randomized clinical trial, a structured moderate-intensity physical activity program reduced the incidence of major mobility disability among sedentary older adults at risk for disability (6). Longitudinal observational studies have also reported lower risks of mobility and functional disability among older adults with higher or increasing levels of physical activity (7–9). However, the LIFE trial evaluated a structured physical activity intervention among older adults already at elevated risk for mobility disability, and observational studies have largely examined differences in overall or changing physical activity levels. Less is known about the functional consequences of initiating regular physical activity in usual life among persistently inactive older adults who are free of mobility, ADL, and IADL difficulties.

A recent study by Wei et al. applied a target trial emulation framework to Health and Retirement Study (HRS) data to examine physical activity initiation and subsequent dementia and cognitive impairment (10). Whether physical activity initiation after persistent inactivity is associated with the subsequent development of new mobility difficulty among older adults who are initially free of functional difficulties remains less clear. Therefore, we conducted a sequential target trial emulation using HRS data to compare physical activity initiators with noninitiators at eligible treatment-decision points. The primary outcome was incident mobility difficulty, with incident ADL and IADL difficulties examined as secondary outcomes.

## METHODS

### Study Population

Data were obtained from the Health and Retirement Study (HRS), which is sponsored by the National Institute on Aging (grant numbers NIA U01AG009740 and NIA R01AG073289) and is conducted by the University of Michigan (11). We used the RAND HRS Longitudinal File 2022 (V1) Documentation file and user guide, which provided detailed information on variable definitions and data processing procedures (12–14). This dataset is a biennial longitudinal cohort from 1992 (Wave 1) to 2022 (Wave 16), with participants periodically replenished approximately every 6 years (3 waves), and included people over age 50 and their spouses, who could be younger than 50 (13,14). Our study focused on older adults.

Physical activity was assessed through two self-reported questions regarding the frequency of moderate and vigorous physical activity, with response options including every day, more than once per week, once per week, one to three times per month, or never (15). In this study, we defined regular physical activity as engaging in moderate or vigorous physical activity at least once per week. Mobility, activities of daily living (ADL), and instrumental activities of daily living (IADL) were used to represent difficulty in daily activities. Mobility included difficulty with walking one block, walking several blocks, climbing several flights of stairs, and climbing one flight of stairs. ADL included difficulty with bathing, eating, dressing, walking across a room, getting in or out of bed, and toileting, whereas IADL included difficulty using the phone, managing money, taking medications, shopping for groceries, and preparing hot meals (12). Mobility, ADL, and IADL were assessed using established HRS measures, with higher scores indicating greater functional difficulty.

For each sequential trial, the baseline was defined as the wave in which eligibility was assessed. Eligibility required that participants had physical activity information available to classify baseline activity initiation status. Additional inclusion criteria required that participants had completed consecutive interviews at baseline and the previous two waves and, at both previous waves, reported not engaging in defined regular physical activity. Participants were also required to be at least 65 years old at the previous wave. Wave 9 was the minimum starting baseline because moderate and vigorous measures were consistently available beginning in Wave 7 of the HRS survey (12,16). Records after Wave 13 were also excluded to ensure that each trial had a theoretical opportunity for 3 subsequent biennial follow-up intervals (approximately 6 years) before Wave 16. We further restricted eligible trial baselines to participants with no mobility, ADL, or IADL difficulties at baseline.

This study used de-identified, public-use HRS data and did not involve direct interaction with study participants or access to identifiable private information. Therefore, institutional review board approval was not required for this secondary analysis.

### Study Design

A sequential target trial emulation design was implemented, specifying the protocol of a hypothetical randomized trial and emulating it using observational data (17). The baseline wave of each eligible trial was considered time zero for follow-up. Treatment assignment was determined at each eligible trial baseline and remained fixed for that trial throughout follow-up. We defined initiators as participants who were inactive at both previous waves and reported regular moderate or vigorous physical activity at baseline, and noninitiators as participants who remained inactive at baseline after being inactive at both previous waves. This treatment definition represented the status of physical activity initiation observed at the baseline interview, rather than the exact time at which physical activity began between the two biennial assessments. Also, a participant could contribute to multiple trial baselines over time. For example, a participant who remained eligible and remained inactive could contribute to multiple noninitiator trials and could subsequently contribute an initiator trial upon reporting regular physical activity. In addition, because some participants may have missed one or more post-baseline follow-up interviews, sequential trials did not imply contribution to every subsequent trial baseline or follow-up wave. Participants who missed a post-baseline interview were censored for that trial, even if they subsequently returned for interview. Eligible trials in which neither post-baseline functional status nor death as a competing event could be ascertained were excluded from the outcome analysis because their post-baseline outcome status could not be determined. **Figure 1** presents a flowchart of the study population selection and sequential trial construction, including eligibility assessment and classification into initiator and noninitiator groups.

**Figure 1.**
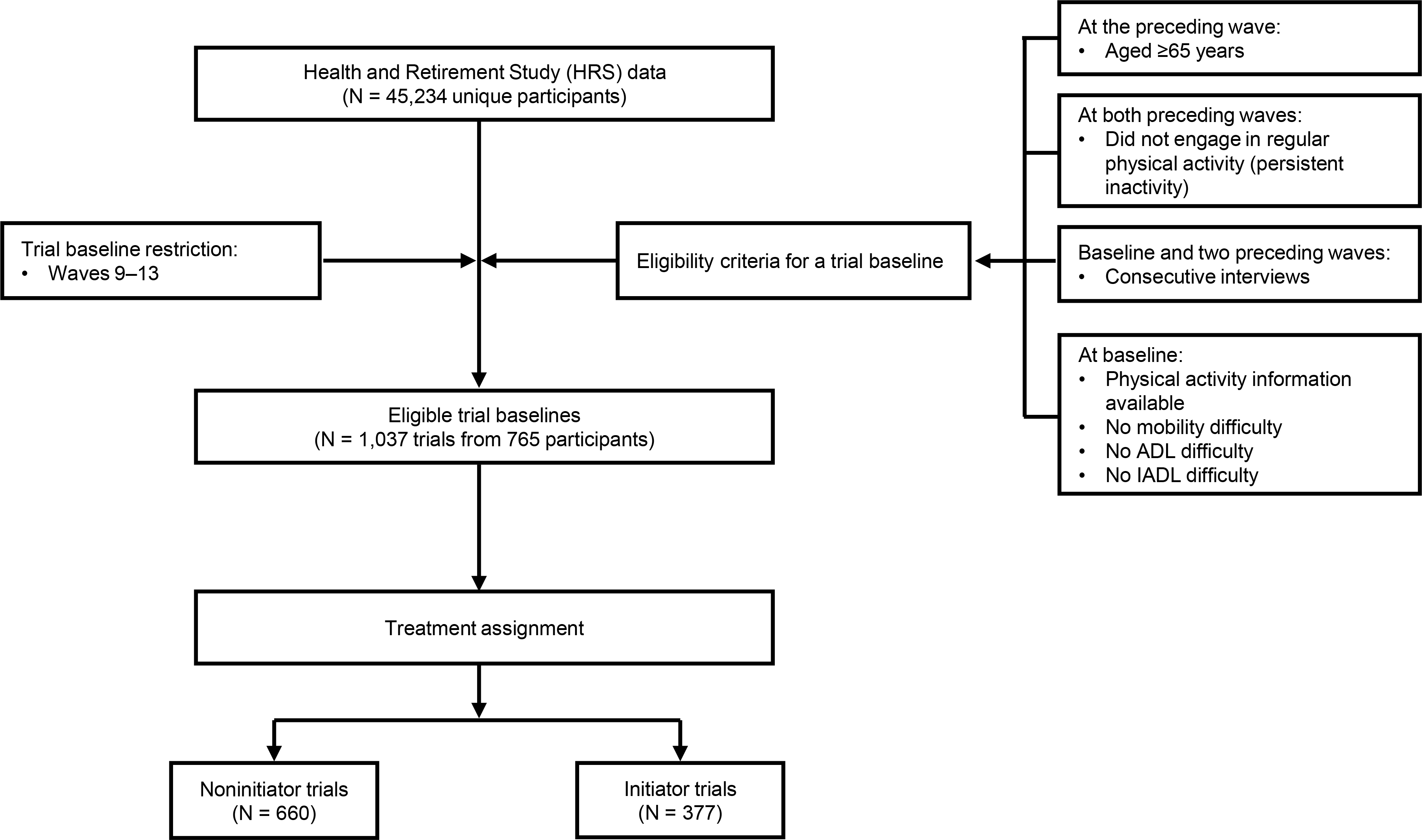
Flowchart of study population selection, eligibility criteria, and sequential target trial construction. Regular physical activity was defined as moderate or vigorous physical activity at least once per week. ADL, activities of daily living; IADL, instrumental activities of daily living; HRS, Health and Retirement Study.

### Outcomes and Covariates

Participants with no difficulty at baseline were considered to have developed incident functional difficulty when they first reported at least one difficulty during follow-up. In this study, the primary outcome was incident mobility difficulty. The secondary outcomes were incident ADL and IADL difficulties, which were examined separately. Death before the incident functional difficulty was treated as a competing event. We also explored individual ADL and IADL components.

To account for potential confounding, we considered pretreatment covariates across multiple domains. Time-fixed covariates included gender, race, ethnicity, and education level. Time-varying covariates from the previous wave were used as pretreatment covariates. These included age at the previous interview, marital status, total household income, total wealth less IRA, self-reported health, ADL/IADL values, mobility, body mass index (BMI), smoking status, eyesight, moderate and vigorous physical activity frequency, and the number of chronic health conditions ever experienced. We also included the change in mobility between the two previous waves to account for past pretreatment functional trajectory. Self-reported health was rated as excellent, very good, good, fair, or poor, and the 8 chronic health conditions were high blood pressure, diabetes, cancer, lung disease, heart disease, stroke, psychiatric problems, and arthritis (12). Given the long time span covered by the survey waves, total household income and wealth were recalculated to 2022 U.S. dollars using the annual average Consumer Price Index for All Urban Consumers (CPI-U) from the U.S. Bureau of Labor Statistics (18). In addition, because total household income and total wealth less IRA were highly skewed, with possible values of 0 or negative for wealth, an inverse hyperbolic sine (IHS) transformation was applied to reduce right skewness (19). Age and IHS-transformed total household income were further modeled using natural cubic splines with 3 degrees of freedom to account for nonlinear relationships and improve covariate balance (20,21). We included the proxy interview factor to account for differences in health and functional status assessments, as well as potential measurement differences associated with respondent type. We also included baseline wave as a categorical covariate to adjust for differences across HRS waves in physical activity initiation and the risk of functional difficulty.

### Statistical Analysis

The distributions of pretreatment covariates were first summarized across eligible trials and compared between treatment groups. Several assumptions were made for causal interpretation of the weighted estimates. These included consistency, positivity, conditional exchangeability given measured pretreatment covariates, and no interference (22). Propensity scores were estimated using multivariable logistic regression, with treatment status (initiators vs noninitiators) as the outcome and the covariates described above used to estimate the probability of physical activity initiation. Stabilized inverse probability of treatment weights (IPTWs) were calculated based on propensity scores to balance observed confounding variables between treatment groups (23). Overlap (positivity) was assessed to determine whether the treatment groups were comparable, and a love plot was generated to display covariate balance using standardized mean differences for continuous variables and differences in proportions for binary variables (24). Absolute differences less than 0.1 indicated adequate covariate balance between treatment groups.

The primary analysis evaluated the average treatment effect (ATE) of initiating regular physical activity on incident mobility difficulty. This effect was expressed as the risk difference in cumulative incidence between treatment groups at approximately 2, 4, and 6 years of follow-up. The estimand was defined as follows:

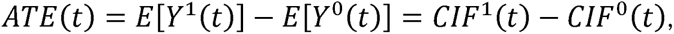

where *CIF*^1^(*t*) and *CIF*^0^(*t*) represent the cumulative incidence of incident mobility difficulty by time *t* among initiators and noninitiators, respectively. IPTW-weighted cumulative incidence functions were estimated using weighted Aalen-Johansen estimators, treating death as a competing event (25–27). Because each participant could contribute to multiple sequential trials, some trials are not independent and are correlated. Therefore, bootstrapping methods were applied to generate 95% confidence intervals, based on 1000 participant-level bootstrap replicates. In each replicate, participants were resampled with replacement together with all sequential trials contributed by each selected participant. The propensity score model and treatment weights were recalculated within each bootstrap replicate.

The secondary analyses examined incident ADL and IADL difficulties separately using the same analytic approach as the primary analysis. Additional analyses examined individual ADL and IADL components to assess whether the overall ADL and IADL findings were driven by specific functional activities.

Three sensitivity analyses were conducted to check the robustness of the primary analysis results. The first sensitivity analysis addressed potentially informative censoring due to nonresponse after baseline. Pooled logistic regression models were fitted to estimate the conditional probability of being uncensored at each follow-up wave (28). The denominator model included treatment assignment, follow-up wave, and pretreatment covariates, while the numerator model included only treatment assignment and follow-up wave. At each follow-up wave, the stabilized censoring weight was calculated as the predicted probability from the numerator model divided by that from the denominator model. These values were multiplied across follow-up waves to obtain the cumulative inverse probability of censoring weight (IPCW) through each follow-up interval. The resulting IPCWs were then multiplied by the stabilized IPTWs from the primary analysis to obtain the final analysis weights (28). In the second sensitivity analysis, we restricted the analysis to the first eligible sequential trial contributed by each participant and repeated the propensity score weighting and outcome analysis. This analysis was conducted to evaluate whether the primary findings were influenced by participants contributing multiple sequential trials. The third sensitivity analysis evaluated the robustness of the estimated treatment effect with respect to the population sampling design by applying HRS combined respondent-level survey weights. Propensity scores were calculated by incorporating survey weights into the logistic regression model. Stabilized IPTWs were then derived from these scores and multiplied by the survey weights to obtain the final analysis weights for estimating the weighted cumulative incidence functions. All analyses were performed using SAS version 9.4 (SAS Institute Inc.) and R version 4.6.1 (R Core Team).

## RESULTS

A total of 1,037 eligible trials were identified, including 660 noninitiator trials and 377 initiator trials. Because participants could contribute to more than one sequential trial, these trials represented 765 unique individuals. **Table 1** summarizes the distribution of characteristics by treatment group. Compared with noninitiators, initiators were slightly younger and had a higher proportion of men. Initiators were also more likely to report moderate or vigorous physical activity one to three times per month, which remained below the study threshold of at least once per week for regular physical activity.

**Table 1.**
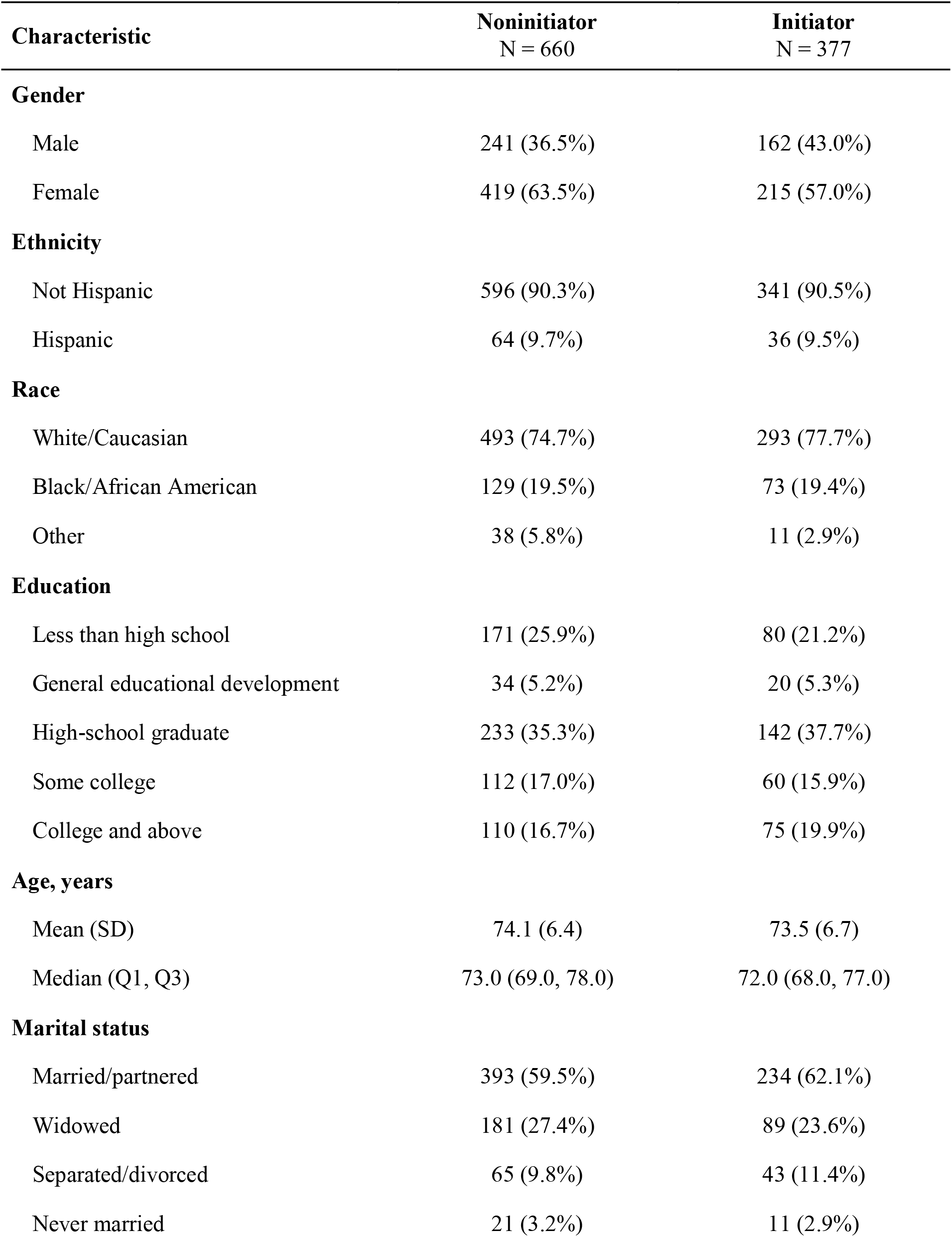
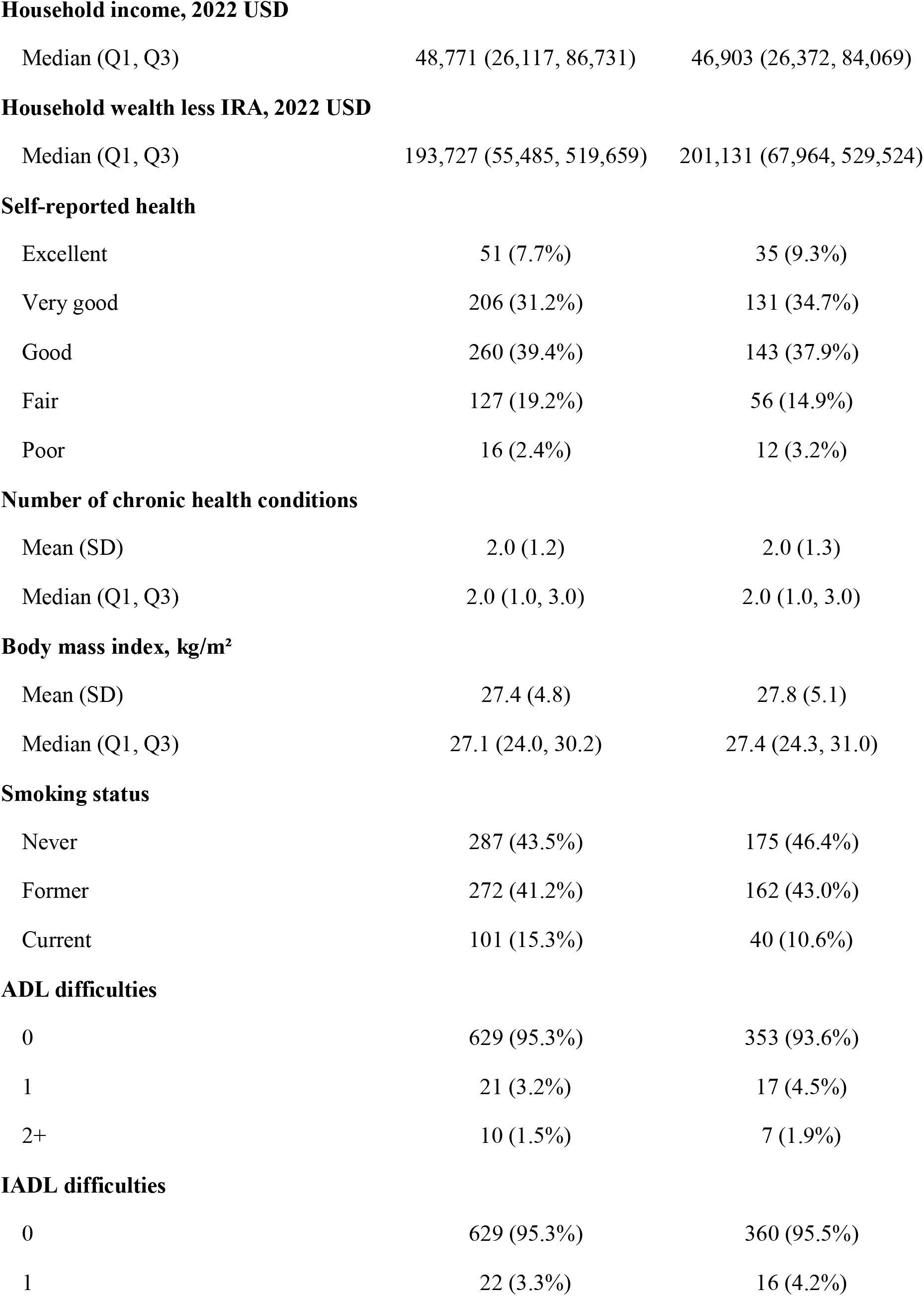
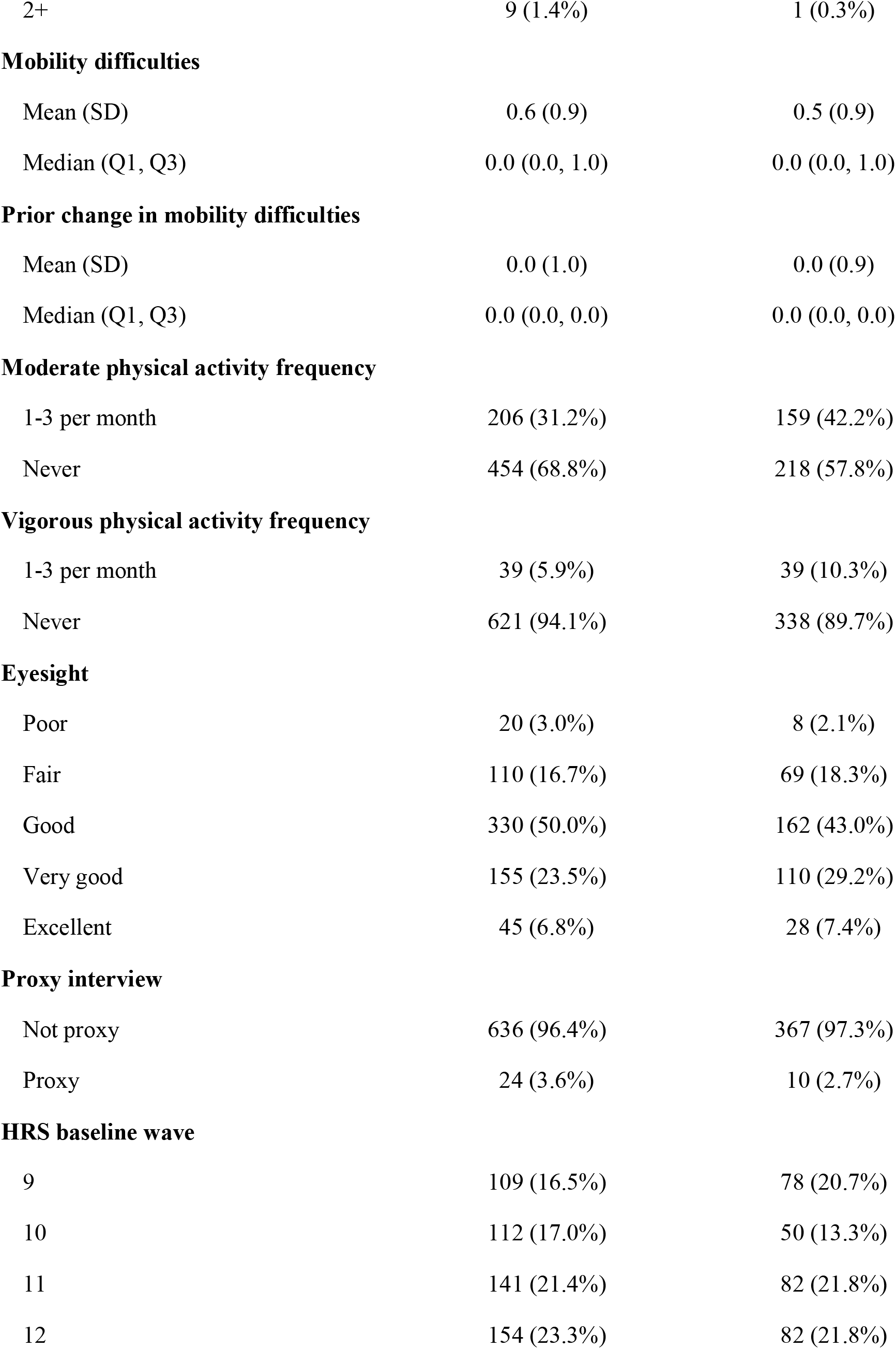
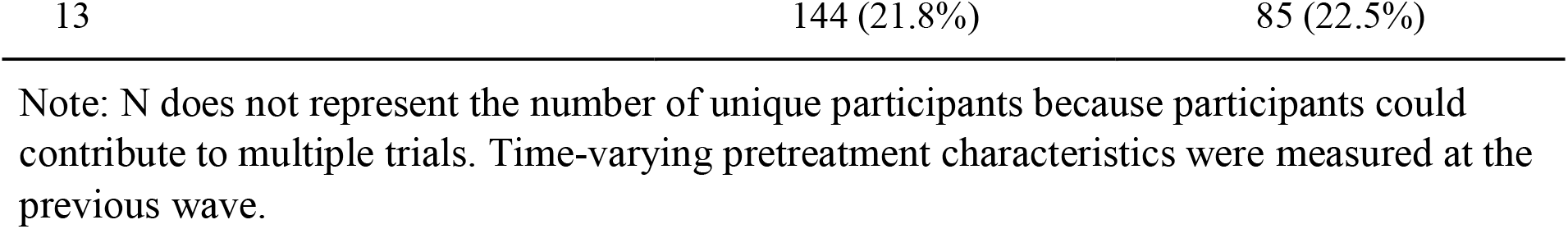
Distribution of Pretreatment Characteristics by Treatment Group.

Propensity score distributions showed good overlap between the treatment groups, and all covariate balance measures were below 0.03, indicating good covariate balance (**Figure 2**). As shown in **Figure 3**, the weighted cumulative incidence of incident mobility difficulty among noninitiators was 39.73%, 53.17%, and 64.96% at approximately 2, 4, and 6 years of follow-up, respectively, compared with 32.75%, 50.27%, and 59.52% for initiators. The risk differences (percentage points; 95% CIs) at 2, 4, and 6 years were −6.98 (−13.11, −0.88), −2.90 (−9.77, 4.29), and −5.44 (−12.27, 1.99), respectively. No clear differences were observed for incident ADL or IADL difficulty at any follow-up time, with all 95% confidence intervals including zero.

**Figure 2.**
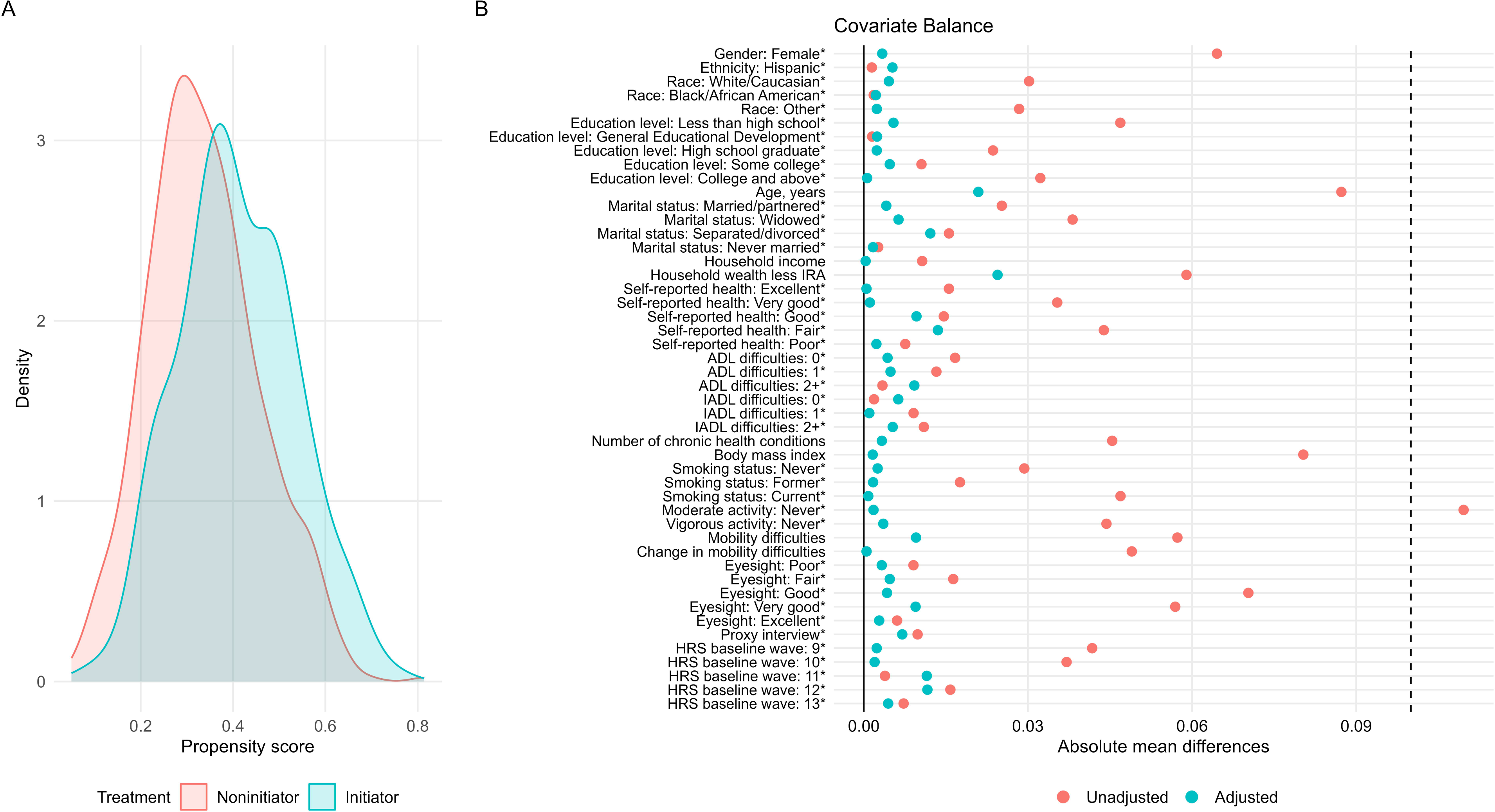
Overlap and balance assessment among eligible trials. Panel A displays the overlap between treatment groups (noninitiator versus initiator). Panel B presents a love plot evaluating covariate balance between treatment groups after application of stabilized inverse probability of treatment weights (IPTWs) derived from the propensity score model. As described in the Methods section, binary variables marked with an asterisk (*) indicate that raw differences in proportions are presented, whereas standardized mean differences are used for continuous variables. Time-varying pretreatment characteristics were measured at the previous wave. The dashed vertical line represents an absolute difference of 0.10, which serves as the threshold for adequate balance. Both unadjusted and adjusted values are shown.

**Figure 3.**
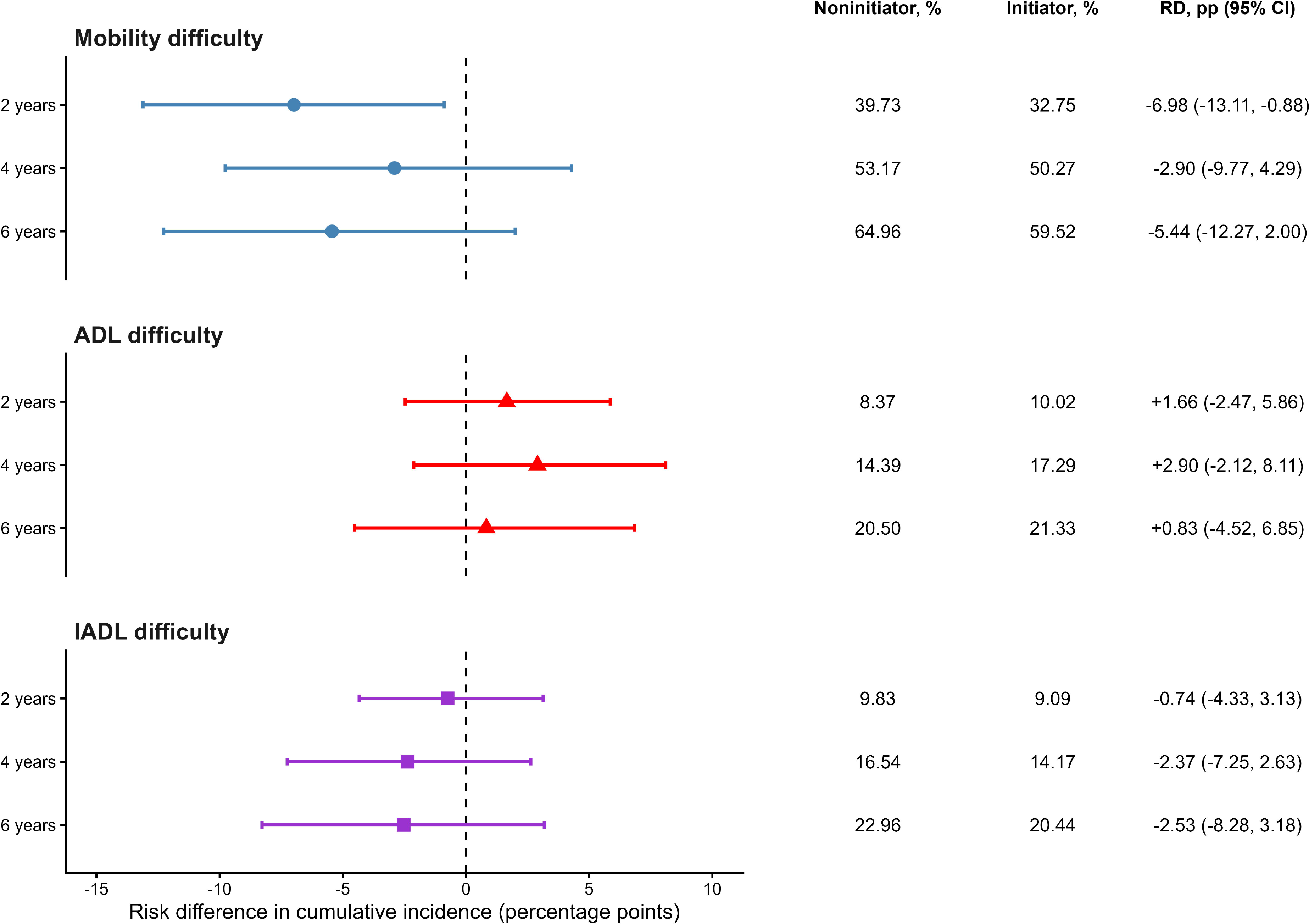
Cumulative incidence and risk differences for incident mobility, ADL, and IADL difficulty by physical activity initiation status. Risk differences were calculated as the cumulative incidence among initiators minus that among noninitiators and are presented in percentage points with participant-level bootstrap 95% confidence intervals at approximately 2, 4, and 6 years. “pp” denotes percentage points. Negative values indicate a lower cumulative incidence among initiators. Risk differences were calculated using unrounded values, which may result in minor discrepancies when subtracting the displayed rounded risks. ADL, activities of daily living; IADL, instrumental activities of daily living; RD, risk difference.

**Figure 4** presents the risk differences (95% CIs) for each ADL and IADL component at the three follow-up times. Estimates varied in direction across individual activities, and all corresponding 95% confidence intervals included zero. Taken together, there was no clear evidence that the overall ADL or IADL findings were driven by a specific activity.

**Figure 4.**
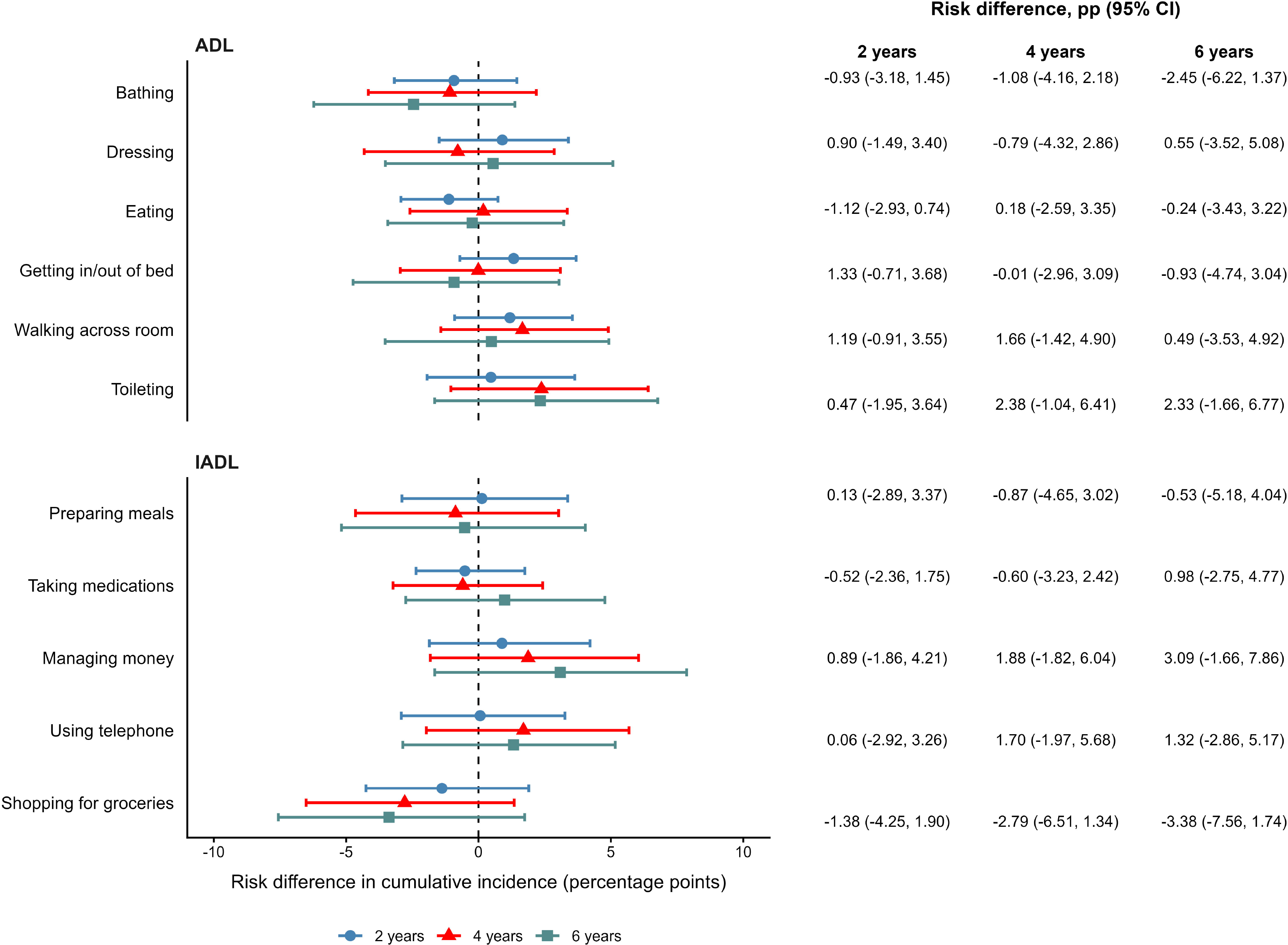
Risk differences for incident difficulty in individual ADL and IADL components by physical activity initiation status. Risk differences were calculated as the cumulative incidence among initiators minus that among noninitiators and are presented in percentage points with participant-level bootstrap 95% confidence intervals at approximately 2, 4, and 6 years. “pp” denotes percentage points. Negative values indicate a lower cumulative incidence among initiators. ADL, activities of daily living; IADL, instrumental activities of daily living; RD, risk difference.

**Table 2** summarizes the sensitivity analysis results. Estimates from the IPCW analysis were very similar to the primary results. For incident mobility difficulty, the risk differences were −7.25 (−13.35, −1.13) at 2 years, −2.78 (−9.99, 4.34) at 4 years, and −5.31 (−12.45, 2.18) at 6 years. Restricting the analysis to the first eligible trial contributed by each participant also produced estimates in a similar direction, although the confidence intervals were wider. The HRS survey-weighted analysis also produced similar estimates, with a 2-year risk difference of −7.51 (−14.45, −0.79), and the estimates at 4 and 6 years remained similar to those from the primary analysis.

**Table 2.** Risk Differences in the Cumulative Incidence of Mobility Difficulty Across Primary and Sensitivity Analyses.

| Analysis | 2-year RD (95% CI) | 4-year RD (95% CI) | 6-year RD (95% CI) |
| --- | --- | --- | --- |
| Primary analysis | -6.98 (-13.11, -0.88) | -2.90 (-9.77, 4.29) | -5.44 (-12.27, 1.99) |
| IPCW <sup>a</sup> | -7.25 (-13.35, -1.13) | -2.78 (-9.99, 4.34) | -5.31 (-12.45, 2.18) |
| First eligible trial only <sup>b</sup> | -6.74 (-13.42, 0.59) | -1.44 (-8.77, 6.43) | -3.41 (-11.05, 4.52) |
| HRS survey-weighted analysis <sup>c</sup> | -7.51 (-14.45, -0.79) | -2.66 (-10.27, 5.31) | -5.49 (-12.99, 2.63) |

Note: Risk differences (RDs) were calculated as the cumulative incidence among initiators minus that among noninitiators and are presented in percentage points with 95% confidence intervals. Negative values indicate a lower cumulative incidence of mobility difficulty among initiators.
a. IPCW denotes inverse probability of censoring weighting.
b. Analysis restricted to the first eligible trial contributed by each participant.
c. Analysis incorporated HRS survey weights.

## DISCUSSION

The purpose of this analysis was to evaluate the effect of initiating regular physical activity on the risk of incident mobility difficulty among older adults with persistent physical inactivity and no mobility, ADL, or IADL difficulties at baseline. Beyond comparing physically active and inactive older adults, we focused on a treatment decision among persistently inactive individuals: whether initiating regular moderate or vigorous physical activity, versus not initiating it, affected subsequent risk of mobility difficulty. Based on the results, at approximately 2 years, physical activity initiation was associated with about a 7-percentage-point lower risk of mobility difficulty. The estimates at 4 and 6 years were also lower among initiators, although they were less precise. We did not observe similar differences for ADL or IADL difficulties. In the sensitivity analyses, the lower 2-year risk was also observed in the IPCW and survey-weighted analyses, with estimates similar in magnitude to the primary analysis. Restricting the analysis to the first eligible trial also produced a comparable estimate, although with wider confidence intervals.

An important strength of this study was the use of a target trial emulation framework. By explicitly defining eligibility requirements, treatment assignment, and the start of follow-up, this approach provides a clearer causal contrast between physical activity initiation and noninitiation and helps avoid potential design biases (17,29). A further methodological strength of this study was the use of a sequential target trial design, in which eligibility and treatment assignment were evaluated at multiple interview waves rather than at a single baseline (17,29,30). For example, a participant could be classified as a noninitiator at one eligible wave and later as an initiator after starting regular physical activity. This better reflects how physical activity changes over time, rather than treating activity status as fixed at a single baseline. In addition, this approach can increase the analytic sample size by allowing more initiator trial entries and may improve the precision of results (29,31). Because trials contributed by the same participant are not independent and could have overlapped follow-up periods, bootstrap resampling was performed at the participant level, retaining all contributed trials for each sampled participant (32,33).

Our results showed a significantly lower incidence of mobility difficulty among physical activity initiators in the short term. This is consistent with findings from the Lifestyle Interventions and Independence for Elders (LIFE) randomized clinical trial, in which a structured moderate-intensity physical activity program reduced the incidence of major mobility disability over an average follow-up of 2.6 years among sedentary older adults at risk for disability (6). Previous longitudinal studies have also found that higher or increasing physical activity was associated with a lower subsequent risk of mobility disability in older adults (7,8,34). Our study builds on this work by focusing on examining the transition from persistent inactivity to regular physical activity, rather than overall activity level, and by evaluating incident mobility difficulty among older adults without functional difficulties at baseline using a sequential target trial emulation.

The difference in mobility difficulty was most apparent at approximately 2 years, while estimates at 4 and 6 years were in the same direction but less precise. One possible explanation is that treatment was defined by physical activity initiation at baseline and did not require participants to maintain the same activity level throughout follow-up. Initiators may later have reduced or stopped regular physical activity, while noninitiators could have become active later but remained classified according to their original trial assignment. Therefore, the estimated effect represents the effect of initiating regular physical activity at an eligible baseline rather than the effect of sustained physical activity over the entire follow-up period (29). Previous studies have also shown that maintaining physical activity over longer periods can be challenging among older adults and that gains in physical activity may diminish over time (35,36). Behavioral changes over time could reduce differences between the treatment groups during longer follow-up.

In contrast to the mobility findings, we observed no clear differences in incident ADL or IADL difficulty between initiators and noninitiators. This pattern is consistent with findings from the LIFE Study, in which a structured physical activity intervention reduced severe mobility disability but did not significantly reduce incident ADL or IADL disability (37). Other intervention studies and systematic reviews have also reported less consistent effects of physical activity on self-reported ADL and IADL outcomes than on physical performance measures (38,39). Mobility may represent a more direct aspect of physical functioning, whereas ADL and IADL difficulties reflect a broader combination of physical, cognitive, medical, and social factors. Additional analyses of individual ADL and IADL activities in our study also did not identify a clear pattern, suggesting that the overall ADL or IADL findings were not driven by a particular activity.

This study contains several limitations. Physical activity was an observational exposure that was not randomized. Although we emulated a target trial framework and adjusted for a broad range of pretreatment demographic, socioeconomic, health, physical activity, and functional characteristics, residual confounding from unmeasured or imperfectly measured characteristics cannot be ruled out. For example, individuals who begin regular physical activity may differ from those who remain inactive in motivation, underlying health, or other factors that are not fully captured in HRS. Physical activity was also self-reported and assessed by frequency category. The HRS measures did not provide information on activity duration or total minutes per week, so our definition of regular physical activity should not be interpreted as meeting recommended physical activity guidelines.

Because HRS physical activity was assessed at biennial interviews, the exact date of initiation was unknown. Some participants classified as initiators may therefore have begun regular activity months before the baseline interview. Since functional status and treatment status were both assessed at that interview, our analysis estimates the effect from the time initiation was observed, rather than from the actual time physical activity began. Similarly, the exact timing of incident functional difficulty between interviews could not be determined. If participants died between interviews, it was not possible to determine whether functional difficulty had occurred before or after death. In addition, many covariates and functional measures were self-reported and may be subject to measurement error.

## CONCLUSION

In conclusion, initiation of regular moderate or vigorous physical activity among persistently inactive older adults without baseline functional difficulties was associated with a lower risk of incident mobility difficulty at approximately 2 years. Estimates at 4 and 6 years were also lower among initiators but were less precise, and we found no clear differences in incident ADL or IADL difficulty. These results suggest a possible benefit of initiating physical activity for mobility, while more detailed data on the timing and persistence of activity are needed to better understand effects at later follow-up.

## CRediT Author Statement

Jingchen Liang: Conceptualization, Data curation, Methodology, Formal analysis, Software, Visualization, Writing - original draft, Writing - review & editing.

## Data Availability

The data used in this study are publicly available from the Health and Retirement Study (HRS) upon registration.

